# Virlaza™ Inhibits Sars-COV-2-induced Inflammatory Response of Bronchial Epithelial Cells and Pulmonary Fibroblast

**DOI:** 10.1101/2021.07.17.21260123

**Authors:** Maysa Alves Rodrigues Brandao-Rangel, Dobroslav Melamed, Anamei Silva-Reis, Boris Brill, Lucas dos Santos Zamarioli, Carlos Rocha Oliveira, Rodolfo P Vieira

## Abstract

Coronavirus disease 2019 (COVID-19), which is currently a global public health emergency and beyond vaccines as a prophylactic treatment, no specific and effective therapeutical treatments are available. COVID-19 induces a massive release of proinflammatory cytokines, which drives COVID-19 progression, severity, and mortality. In addition, bronchial epithelial cells are the first pulmonary cells activated by coronavirus-2 (SARS-Cov-2) leading to massive cytokine release, which can hyperactivate lung fibroblasts, resulting in pulmonary fibrosis, a phenomenon observed even in moderate COVID-19 survivors. This in vitro study tested the hypothesis that Virlaza^™^, a herbal medicine, could inhibit the hyperactivation of human bronchial epithelial cells (BEAS-2B) and pulmonary fibroblasts (MRC-5) induced by SARS-Cov-2. BEAS-2B (5×10^4^/mL/well) and MRC-5 (5×10^4^/mL/well) cells were co-cultivated with 1ml of blood of a Sars-Cov-2 infected patient for 4 hours and Virlaza^™^ (1ug/mL) was added in the first minute of the co-culture. After 4 hours, the cells were recovered and used for analysis of cytotoxicity by MTT and for mRNA expression of P2X7 receptor E iNOS. The supernatant was used to measure ATP and cytokines. Sars-Cov-2 incubation resulted in increased release of ATP, IL-1beta, IL-6, IL-8, and TNF-alpha by BEAS-2B and MRC-5 cells (p<0.001). Treatment with Virlaza^™^ resulted in reduction of ATP, IL-1beta, IL-6, IL-8, and TNF-alpha release (p<0.001). In addition, Sars-Cov-2 incubation resulted in increased expression of P2X7 receptor and iNOS (p<0.001), which has been reversed by Virlaza^™^ (p<0.001). In conclusion, Virlaza^™^ presents important anti-inflammatory effects in the context of Sars-Cov-2 infection.

## Introduction

The quick spread of Coronavirus disease (COVID-19) resulted in a pandemic in the whole world, reaching more than 100 million cases and 2 million deaths due to the severe acute respiratory syndrome coronavirus 2 (SARS-CoV-2) [1]. The progression, severity, and mortality of COVID-19 differ from no, mild, or moderate symptoms in most of the patients, while up to 15% develop the severe and critical form of the disease with high mortality rates [2]. Among the factors underlying the development of the severe and critical forms of COVID-19, advanced age [3], comorbidities [3], hyperactivation of the immune system [3,4] are proven to have a central role, hyperactivation of the immune system results in the cytokine storm, which is characterized by the synthesis and release of high levels of proinflammatory cytokines [3,4]. In this way, cytokine storm plays a major role in Covid-19 pathogenesis [3,4].

In this context, bronchial epithelial cells have been described not only as a mechanical barrier for the respiratory tract, but also as an important cell layer centrally involved in the innate immune response, which actively synthetize and release a plethora of inflammatory and fibrotic mediators [5], capable to activate fibroblasts inducing fibrosis. In addition, activation of the purinergic receptor P2X7 in the airway epithelium has been described as a key receptor involved in the inflammatory [6] and fibrotic [7] responses of the lungs to a variety of stimulus, including SARS-CoV-2 [8]. Furthermore, P2X7 is also involved in pulmonary fibroblast proliferation and activation, driven the pulmonary fibrotic response as well [7].

Activation of P2X7 receptor may be a trigger by increased levels of nitric oxide (NO) mediated by increased expression and activation of inducible nitric oxide synthase (iNOS) [9]. Indeed, increased iNOS expression drives an increased inflammatory and fibrotic responses of the lungs, such as in asthma [10], pulmonary fibrosis [11], and bacterial infections [12, 13].

Thus, this study tested the hypothesis that Virlaza^™^ could inhibit bronchial epithelial cells and lung fibroblast activation in vitro, involving iNOS and P2X7 signaling.

## Materials and Methods

This study and all experimental procedures were analyzed and approved by ethical committee of School of Medicine of Anhembi Morumbi University (4.637.625 in 08 of April of 2021) and were carried out in accordance with Declaration of Helsinki for ethical principles for medical research involving human subjects.

### Human Cell Co-Culture Study

Virlaza^™^ [Clove glycerin extract (1:5); Eucalyptus Glycerin Extract (1:5); Basil Glycerin Extract (1:5); Sage Glycerin Extract (1:5); Maritime pine Glycerin extract (1:5); Clove Tincture (ethyl alcohol 99.8%) (1:5); Eucalyptus Tincture (1:5); Maritime pine Tincture (ethyl alcohol 99.8%) (1:5); Basilica Tincture (ethyl alcohol 99.8%); Sage Tincture (ethyl alcohol 99.8%) (1:5); Selenite (0.0051%), and Zinc (0.32%).

Human bronchial epithelial cell line (BEAS-2B) and human lung fibroblast cell line (MRC-5), which are commercially available of immortalized cell lines, were co-cultured with whole blood from an infected patient with Sars-Cov-2. BEAS-2B and MRC-5 cells were obtained from Rio de Janeiro Cell Bank and co-cultured in DMEM high glucose medium (Sigma Chemical Co., St. Louis, MO, USA) supplemented with 10% fetal calf serum in 500ul of whole blood of Sars-Cov-2 infected male (35-40) years old patient (original Wuhan Sars-Cov-2), presenting an estimated high viral load based on the cycle threshold (CT = 15). Both BEAS-2B and MRC-5 cells were co-cultured at a concentration of 5×10^4^/mL/well in 48 wells plate, at a humidified atmosphere in a CO2 incubator (5% CO2, 37°C) [14]. The blood was collected immediately after hospitalization, prior any antibiotic and corticosteroid administration. The written consent inform was obtained from the patient volunteer for this study. The cells (BEAS-2B or MRC-5) were incubated at the same time with Virlaza^™^ (1ug/mL) and with 500ul of whole blood of Sars-Cov-2 infected patients and the cells and supernatant was obtained 4 hours after stimulation. The cells and supernatant were recovered, centrifuged at 900g for 5 minutes at 4°C, and then the supernatant immediately used for ATP measurement and later for cytokine measurement. The cells were washed using phosphate-buffered saline (PBS) and then subjected to RT-PCR protocol.

### Cytotoxicity assay by MTT assay

To assess the Virlaza^™^ cytotoxicity through cell viability, MTT assay was carried out. Briefly, 5×10^4^ viable BEAS-2B and MRC-5 cells were placed into clear 96-well flat-bottom plates (Corning USA) in DMEM high glucose medium supplemented with 10% fetal calf serum and immediately after different concentrations (0.1ug/mL; 1ug/mL; 10ug/mL; 100ug/mL; and 1000ug/mL) of Virlaza^™^. Following 24h after incubation in a humidified atmosphere of a CO2 incubator (5% CO2, 37°C), 10μL/well of MTT (5mg/mL) was added to the cells (both in the control and Virlaza^™^), which was incubated for 4h. After this time, 100μL of 10% sodium dodecyl sulfate (SDS) solution in deionized water was added to the cells and incubated overnight [15, 16]. The absorbance was measured at 595nm in a benchtop multimode reader SpectraMax i3 (Molecular Devices, San Jose, CA, USA).

### Adenosine Triphosphate (ATP) Measurement

The ATP concentration was determined in the cell culture supernatant immediately after its collection by using the ATPlite Luminescence Assay System (Perkin Elmer, Waltham, MA, USA), according to the manufacturer’s instructions. The results were expressed in nmol/mL [17].

### Cytokines Measurement

The levels of interleukin (IL)-1beta (DY201), IL-1RA (DY280), IL-6 (DY206), IL-8 (DY208) and TNF-alpha (DY210) were measured in the cell culture supernatant by using DuoSet ELISA kit (R&D Systems; Minneapolis, MN, USA) according to the manufacturer’s recommendations using the SpectraMax i3 multiplate reader (Molecular Devices, CA, USA). The results were expressed as pg/mL [14-17].

### Reverse transcriptase-polymerase chain reaction (RT-PCR)

Total RNA isolation from cell pellets was used RNeasy mini kits (Qiagen, Hilden, Germany). Reverse transcription was used Stratascript reverse transcriptase (Stratagene, CA, USA) and random primers (Invitrogen, Germany). Quantitative PCR used Taqman Universal PCR Mastermix (Applied Biosystems, USA) and preformulated primers and probe mixes (Assay on Demand, Applied Biosystems, USA). PCR conditions were 2 min at 50°C, 10 min at 95°C, followed by 45 cycles of 15 s at 95°C and 60°C for 1 min using the CFX96 thermal cycler (Bio rad, Hercules CA, USA). PCR amplification of the housekeeping gene encoding Glyceraldehyde-3-phosphate dehydrogenase (GADPH) was performed during each run for each sample to allow normalization between samples. β-actin was used as a control and correction factor for the expression of the P2X7 receptor, for which the sequences of primers are described as follows: β-actin - forward (5′GTGGGCCGCTCTAGGCACCA3′) and reverse primers (5′CTCTTTGATGTCACGCACGATTTC3′ 540 bp) [15] and P2X7 - forward, 5′ AGATCGTGGAGAATGGAGTG 3′, and (5′ TTCTCGTGGTGTAGTTGTGG 3′) reverse primers [15]. For iNOS – forward 5′-CTATCAGGAAGAAATGCAGGAGAT-3′, and Reverse 5′-GAGCACGCTGAGTACCTCATT-3′ primers [16].

### Statistical Analysis

The software Graph Pad Prism 5.0 was used to perform the statistical analysis and build de graphs. The results were expressed as mean ± standard□error□of□the mean□(SEM) from at least three independent experiments. One-way analysis of variance (ANOVA) was used for multiple comparisons, followed by the Bonferroni post hoc test for comparison among groups. A p-value <0.05 was considered significant.

## Results

### Effects of Virlaza^™^ on ATP Release in BEAS-2B and MRC-5 Cells

Figure 1 shows that Sars-Cov-2 activated BEAS-2B and MRC-5 cells, resulting in increased release of ATP by BEAS-2B (Figure 1, A; p<0.001) and by MRC-5 (Figure 1 B; p<0.001) cells, which has been significantly attenuated by Virlaza^™^, both in BEAS-2B (Figure 1 A; p<0.001) and by MRC-5 (Figure 1 B; p<0.001) cells.

**Figure 1.**
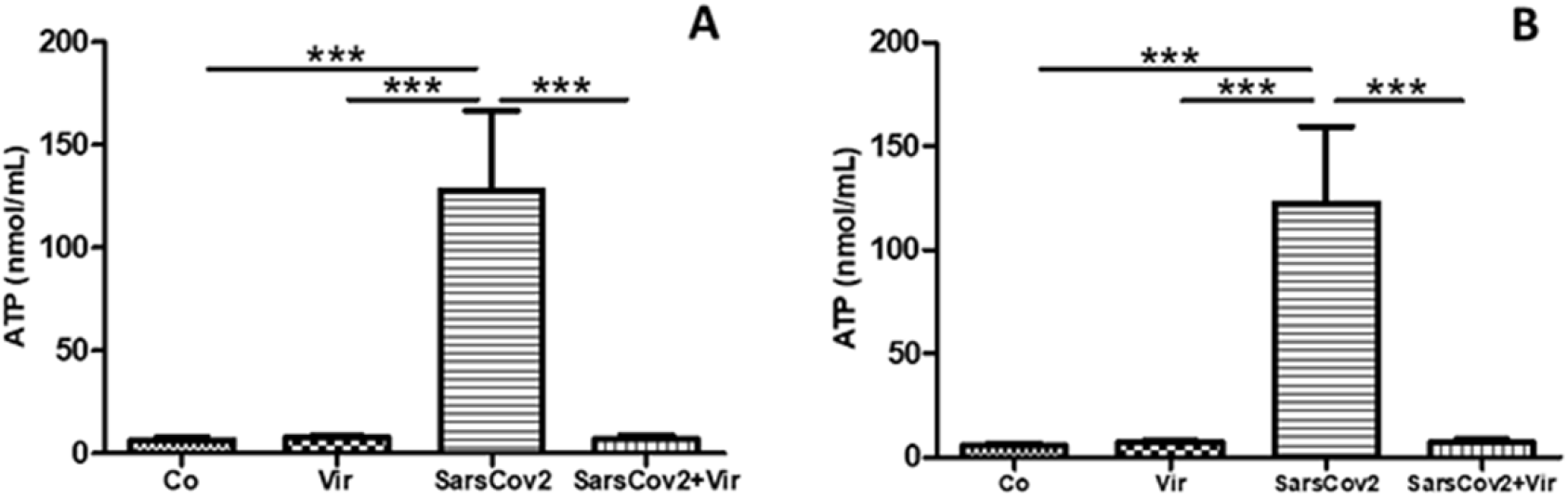
Effects of Virlaza^™^ on ATP Release in BEAS-2B and MRC-5 Cells. Adenosine triphosphate (ATP) levels in the supernatant of the cells. Figure 1 A correspond to experiments done in BEAS-2B cells. Figure 1 A correspond to experiments done in MRC-5 cells. ^***^ p<0.0001. Co = Control; Vir = Treated with 10ug/mL of Virlaza^™^; SarsCov2 = Stimulated with blood infected with Sars-Cov-2; SarsCov2+Vir = Stimulated with blood infected with Sars-Cov- 2 + 10ug/mL of Virlaza^™^.

### Effects of Virlaza^™^ on Bronchial Epithelial Cells (BEAS-2B) Activation

Figure 2 shows that Sars-Cov-2 activation resulted in increased synthesis and release of IL-1beta (Figure 2, A; p<0.001), IL-6 (Figure 2 C; p<0.001), IL-8 (Figure 2 D; p<0.001), and TNF-alpha (Figure 2 E; p<0.001), while Virlaza^™^ resulted in a significant reduction of the synthesis and release of all these pro-inflammatory cytokines (p<0.001). On the other hand, on Sars-Cov-2 stimulated cells, Virlaza^™^ resulted in a significant increase in the synthesis and release of IL-1RA (Figure 2 B; p<0.001), a potent anti-inflammatory cytokine.

**Figure 2.**
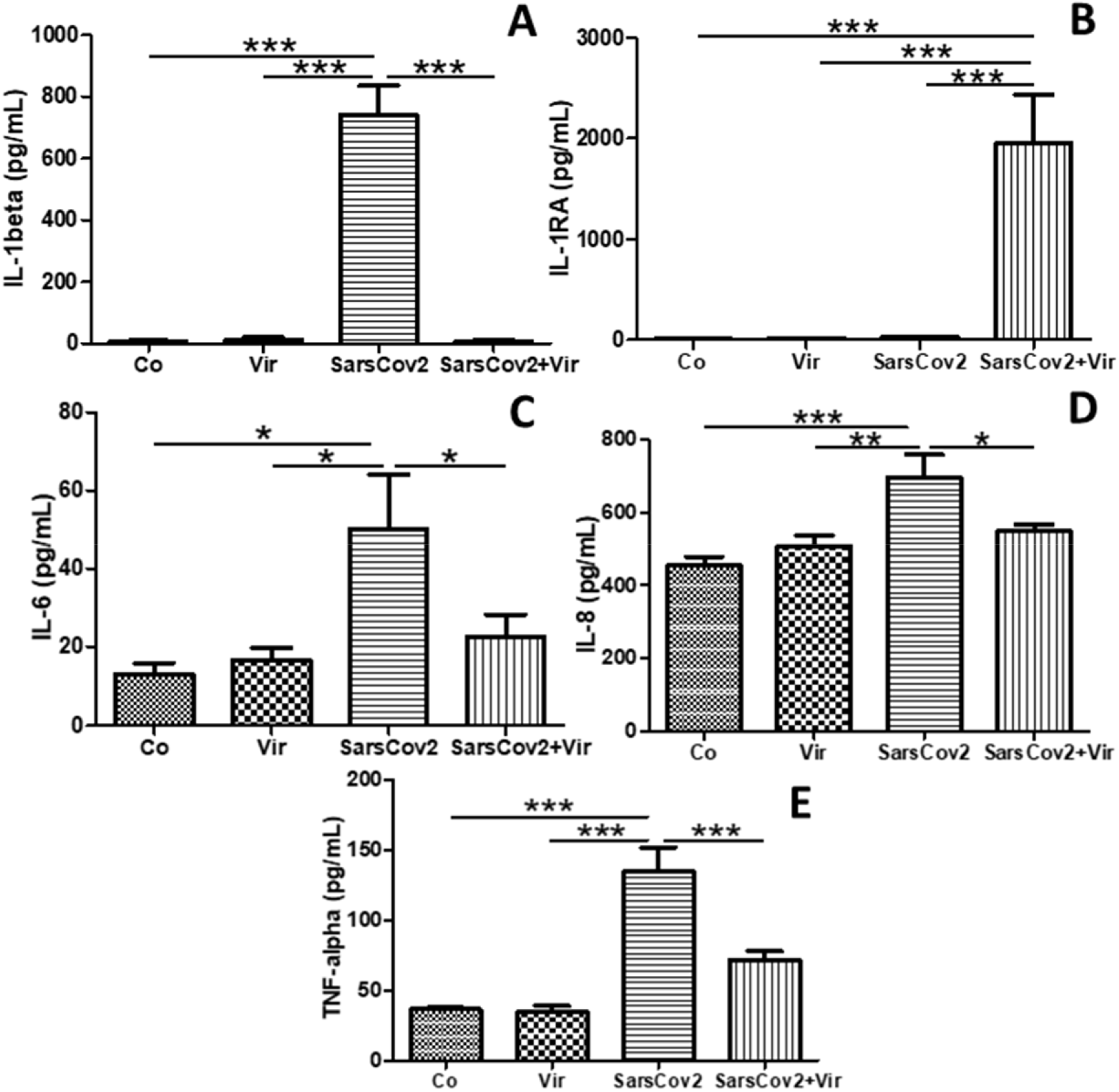
Effects of Virlaza^™^ on Bronchial Epithelial Cells (BEAS-2B) Activation. Cytokines levels in the supernatant of BEAS-2B cells. ^*^p<0.05; ^**^p<0.01; ^***^ p<0.0001. Co = Control; Vir = Treated with 10ug/mL of Virlaza^™^; SarsCov2 = Stimulated with blood infected with Sars-Cov-2; SarsCov2+Vir = Stimulated with blood infected with Sars-Cov-2 + 10ug/mL of Virlaza^™^.

### Effects of Virlaza^™^ on Pulmonary Fibroblasts (MRC-5) Activation

Figure 3 shows that Sars-Cov-2 activation resulted in increased synthesis and release of IL-1beta (Figure 3 A; p<0.001), IL-6 (Figure 3 C; p<0.001), and TNF-alpha (Figure 3 E; p<0.001), but not of IL-8 (Figure 3 D; p>0;05), while Virlaza^™^ resulted in a significant reduction of the synthesis and release of all these pro-inflammatory cytokines (p<0.001). On the other hand, in Sars-Cov-2 stimulated cells, Virlaza^™^ resulted in a significant increase in the synthesis and release of IL-1RA (Figure 3 B; p<0.001), a potent anti-inflammatory cytokine.

**Figure 3.**
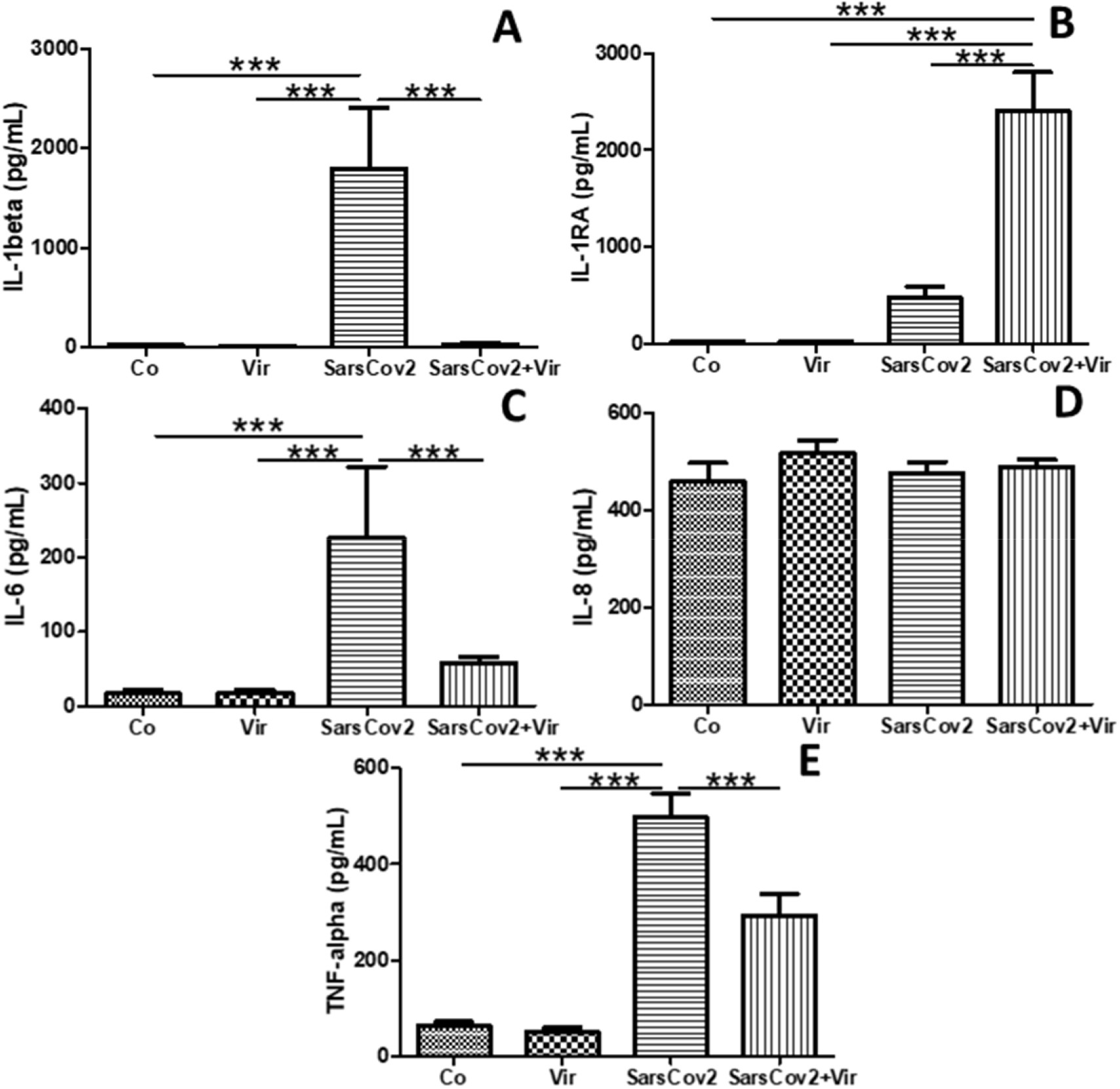
Effects of Virlaza^™^ on Pulmonary Fibroblasts (MRC-5) Activation. Cytokines levels in the supernatant of MRC-5 cells. ^***^ p<0.0001. Co = Control; Vir = Treated with 10ug/mL of Virlaza^™^; SarsCov2 = Stimulated with blood infected with Sars-Cov-2; SarsCov2+Vir = Stimulated with blood infected with Sars-Cov-2 + 10ug/mL of Virlaza^™^.

### Effects of Virlaza^™^ on P2X7 receptor expression in BEAS-2B and MRC-5 Cells

Figure 4 shows that Sars-Cov-2 activation resulted in increased expression of P2X7 receptor in BEAS-2B (Figure 4 A; p<0.001), and in MRC-5 (Figure 4 B; p<0.001), while Virlaza^™^ resulted in a significant reduction of the expression of P2X7 receptor in BEAS-2B (Figure 4 A; p<0.001), and in MRC-5 (Figure 4 B; p<0.001) cells.

**Figure 4.**
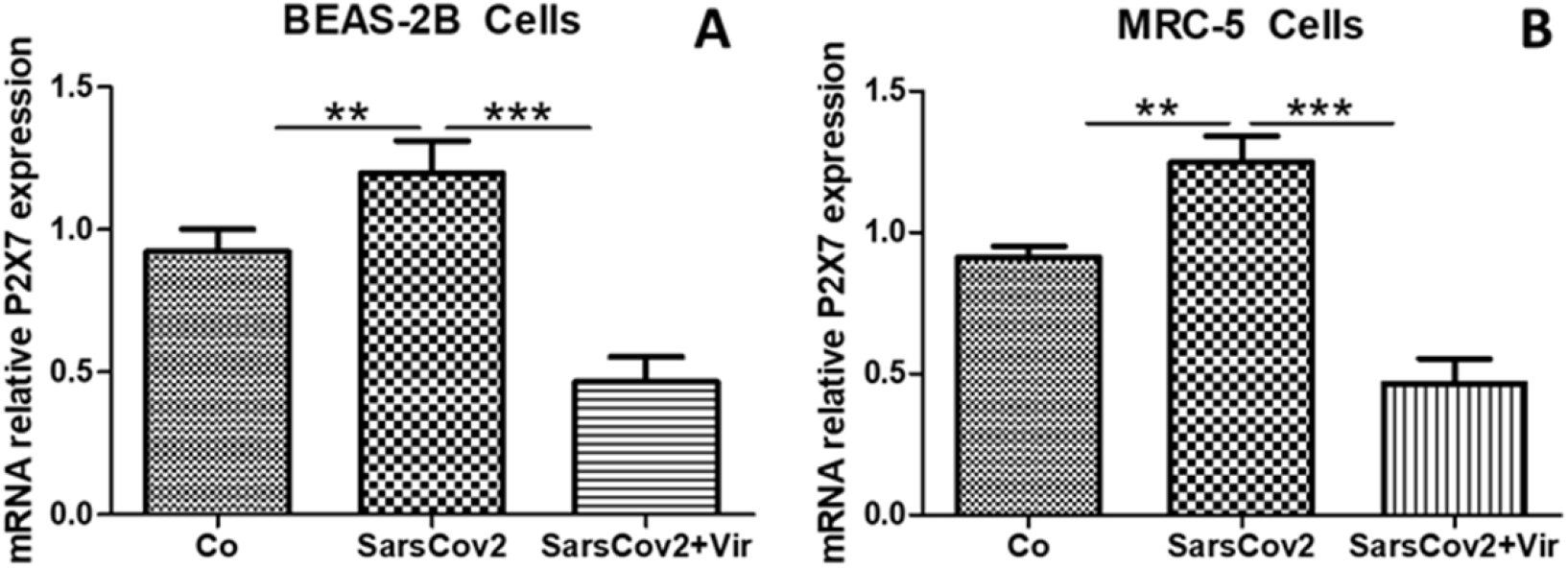
Effects of Virlaza^™^ on P2X7 receptor expression in BEAS-2B and MRC-5 Cells. P2X7 Receptor mRNA expression in BEAS-2B and MRC-5 cells. ^**^p<0.01; ^***^ p<0.0001. Co = Control; Vir = Treated with 10ug/mL of Virlaza^™^; SarsCov2 = Stimulated with blood infected with Sars-Cov-2; SarsCov2+Vir = Stimulated with blood infected with Sars-Cov-2 + 10ug/mL of Virlaza^™^.

### Effects of Virlaza^™^ on iNOS mRNA Expression in BEAS-2B and MRC-5 Cells

Figure 5 shows that Sars-Cov-2 activation resulted in increased expression of iNOS in BEAS-2B (Figure 5 A; p<0.001), and in MRC-5 (Figure 5 B; p<0.001), while Virlaza^™^ resulted in a significant reduction of the expression of iNOS in BEAS-2B (Figure 5 A; p<0.001), and in MRC-5 (Figure 5 B; p<0.001) cells.

**Figure 5.**
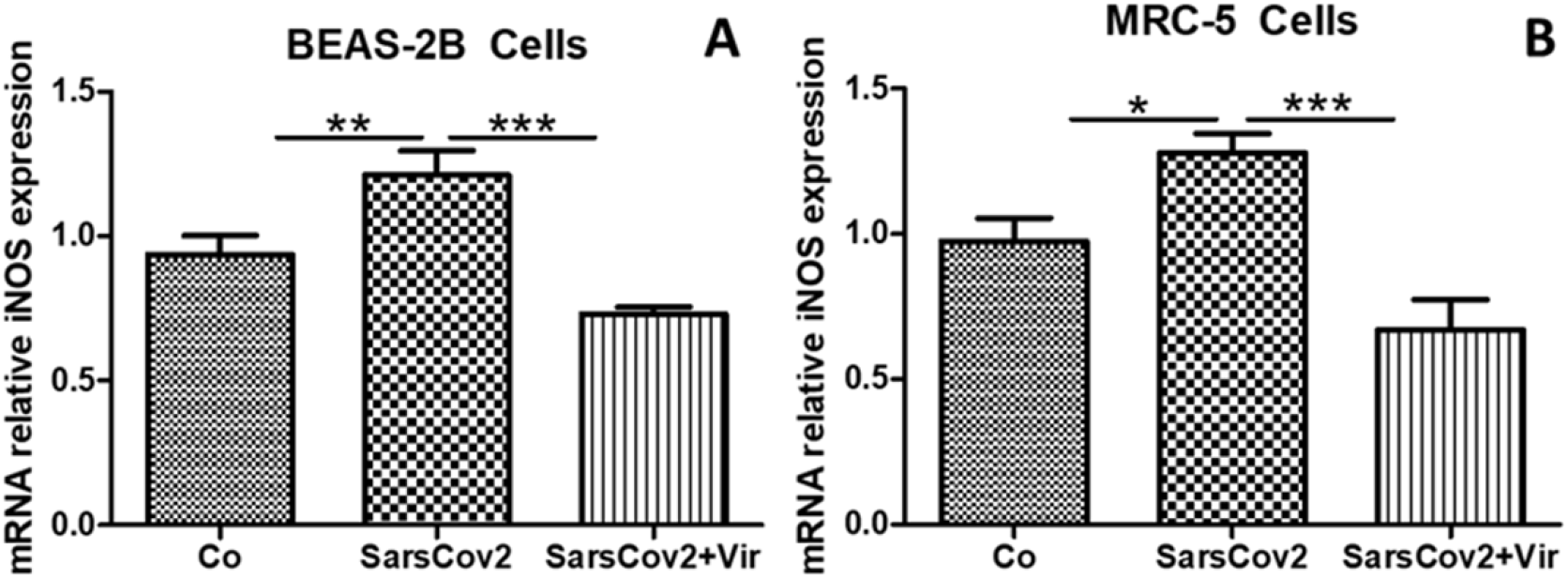
Effects of Virlaza^™^ on iNOS mRNA Expression in BEAS-2B and MRC-5 Cells. P2X7 Receptor mRNA expression in BEAS-2B and MRC-5 cells. ^*^p<0.05; ^**^p<0.01; ^***^ p<0.0001. Co = Control; Vir = Treated with 10ug/mL of Virlaza^™^; SarsCov2 = Stimulated with blood infected with Sars-Cov-2; SarsCov2+Vir = Stimulated with blood infected with Sars-Cov-2 + 10ug/mL of Virlaza^™^.

### Effects of Virlaza^™^ on Cell Viability of BEAS-2B and MRC-5 Cells

Figure 6 shows the different concentrations of Virlaza^™^ on cell toxicity to determine the IC50 value. Figures 6 A (BEAS-2B cells) and 6 B (MRC-5 cells) show that 10 μg/mL was reached, was the dose corresponding to IC50, and was chosen was the study dose.

**Figure 6.**
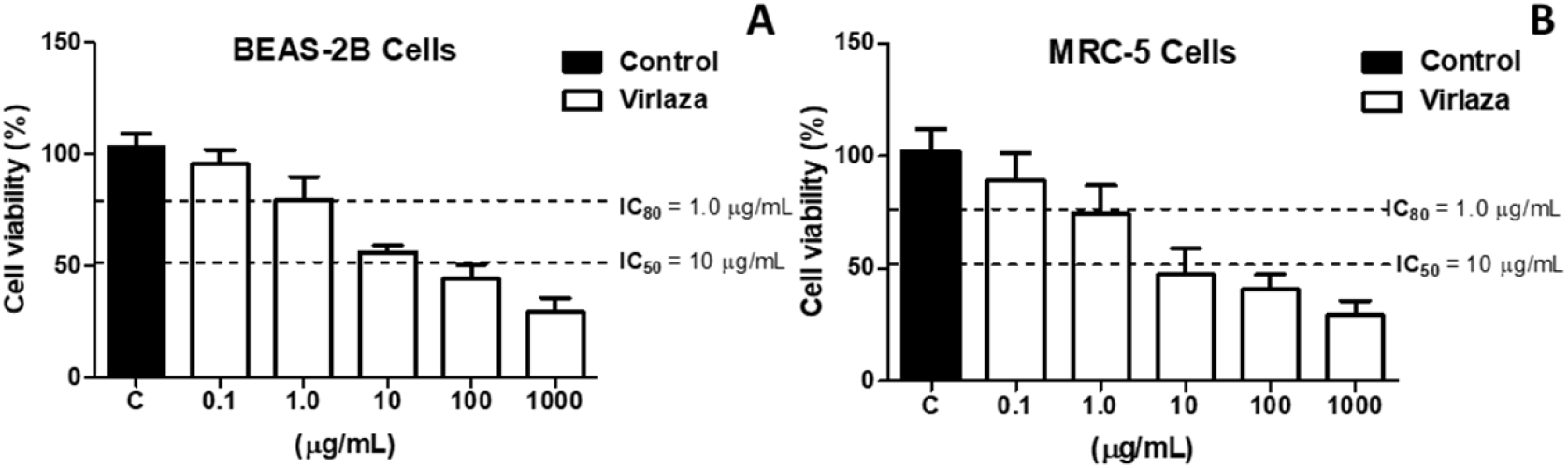
Effects of Virlaza^™^ on Cell Viability of BEAS-2B and MRC-5 Cells. Cell viability (%) measured by MMT assay in BEAS-2B and MRC-5 cells stimulated with growing doses of Virlaza^™^ (0.1ug/mL, 1ug/mL, 10ug/mL, 100ug/mL, 1000ug/mL).

## Discussion

This study shows for the first time that Virlaza^™^ was able to block the cytokine storm caused by Sars-Cov-2-induced epithelial (BEAS-2B) and lung fibroblast (MRC-5) activation, involving dampening of exacerbated purinergic signaling and iNOS expression. In addition, the study demonstrated the safety of an active therapeutic dose Virlaza^™^ in the context of cellular toxicity.

Considering the lack of effective treatment for COVID-19, and the evidence that severely ill COVID-19 patients develop a cytokine storm, some studies are testing the hypothesis that the blockade of some proinflammatory cytokines, such as IL-6 [18] and IL-1beta [19] could be beneficial for severely ill COVID-19 patients. In addition, beyond massive infiltration by neutrophils and macrophages into the lungs, increased blood levels of IL-1β, IL-6, and TNF-alpha have been linked to higher severity and mortality in COVID-19 patients [18, 19]. In this context, bronchial epithelial cells are the main entrance door and the main target of different types of respiratory bacteria and viruses, harming the immune system by exacerbating release of pro-inflammatory cytokines and growth factors [20, 21]. In the present study, we demonstrated that bronchial epithelial cells (BEAS-2B) when stimulated with Sars-Cov-2, responded to increased release of IL-1beta, IL-6, IL-8 and TNF-alpha, which were abolished by treatment with Virlaza^™^, clearly demonstrating the efficacy of Virlaza^™^ blocking the epithelial hyperactivation in the context of Sars-Cov-2.

Importantly, the literature demonstrates the importance of increased expression of P2X7 receptor and consequent activation triggering proinflammatory and pro-fibrotic response in the context of respiratory diseases [4-7]. Such importance has been demonstrated in several cells of the hematopoietic system [6], but also in lung structural cells, such as in epithelial cells [4, 5] and lung fibroblasts [7]., the activation of P2X7 receptors is classically related to the initiation, maintenance, progression, and severity of inflammatory states triggering IL-1beta release [7-9]. Herein, this study demonstrated that Virlaza^™^ inhibited IL-1beta release not only by epithelial cells (BEAS-2B) but also by lung fibroblasts (MRC-5). Particularly about fibroblasts, the inhibitory effect of Virlaza^™^ inhibiting the release of IL-1beta is of particular importance, since IL-1beta directly stimulates collagen synthesis and proliferation in fibroblasts [22, 23]. Therefore, it is plausible to hypothesize that Virlaza^™^ could also have a potential effect to inhibit lung fibrosis, which is commonly observed in COVID-19 survivors [24]. Such observed effects of Virlaza^™^ on IL-1beta release can be supported by the additional results of the present study, which demonstrated that Virlaza^™^ inhibited ATP release and accumulation, resulting in decreased expression of P2X7 receptor in BEAS-2B and in MRC-5 cells. Therefore, further clinical trial investigating the effects of Virlaza^™^ in COVID-19 survivors were guaranteed.

Beyond P2X7 receptor, inducible nitric oxide synthase (iNOS), is though as a key enzyme involved in all aspects of the pathophysiological process of pulmonary infections with diverse etiologies, including for the Sars-Cov-2 [25, 26]. In the context of COVID-19, intense and diffuse pulmonary inflammation results in endothelial dysfunction of the lung vasculature, uncoupling eNOS activity, lowering nitric oxide (NO) production, resulting in different pulmonary alterations and coagulopathy [25, 26]. On the contrary, viral infections trigger an increasing in iNOS activity, which may be acutely advantageous for host defense, once NO plays antiviral effects. However, sustained overproduction of NO mediates deleterious proinflammatory effects [25, 26]. In the present study, it was observed that Sars-Cov-2 increased iNOS expression in epithelial cells (BEAS-2B) and in lung fibroblasts (MRC-5 cells), which was dampened by Virlaza^™^. Interestingly, the literature demonstrates that NO synthesized specifically by the airway epithelium is vital to antiviral, inflammatory, and immune defense of the lungs [26]. However, again, the increased NO mediated by iNOS is related to an unresolved inflammatory process [27] as well as pro-fibrotic response [28]. iNOS overexpression is linked to the release of proinflammatory cytokines, such as IL-1beta, IL-6, IL-8, and TNF-alpha, particularly in the context of acute respiratory distress syndrome (ARDS), phenomena present in cases of severe COVID-19 [27]. In the present study, Sars-Cov-2 stimulation resulted in an increase expression of iNOS by epithelial cells (BEAS-2B) and in lung fibroblasts (MRC-5 cells) and increased release of IL-1beta, IL-6, IL-8, and TNF-alpha, Virlaza^™^ significantly reduced iNOS expression and pro-inflammatory cytokines release, reinforcing the anti-inflammatory and anti-fibrotic effects of Virlaza^™^. These findings concerning the iNOS inhibition by Virlaza^™^ can be strengthened once the literature already points out the possible therapeutic role of iNOS blockade for acute and long COVID-19 [29].

Beyond to inhibit the release of proinflammatory cytokines (IL-1beta, IL-6, IL-8, and TNF-alpha), Virlaza^™^ also induced the release of anti-inflammatory cytokine IL-1RA by epithelial cells (BEAS-2B) and by lung fibroblasts (MRC-5 cells). Functionally, interleukin-1 receptor antagonist (IL-1RA) inhibits the activities of interleukin 1 alpha (IL-1alpha) and interleukin 1 beta (IL-1beta), modulating a wide range of immune and inflammatory responses driven by interleukin 1 related cytokines [30]. Anakinra is a recombinant IL-1RA that has been tested for the treatment of COVID-19 patients, resulting in better clinical outcomes [31-33].

Therefore, the present study concludes that Virlaza^™^ was capable not only to inhibit the release of proinflammatory cytokines but also to stimulate the release of the anti-inflammatory cytokine IL-1RA, which has already been demonstrated to be a promising therapeutic option to prevent the development of severe forms of COVID-19 including respiratory failure, as well to treat the severe forms of COVID-19, improving the clinical outcomes and survival [31-33]. Thus, based on the findings of the present study, further clinical trials with Virlaza^™^ for the prevention and treatment of COVID-19 are guaranteed.

## Data Availability

The raw data will be available upon a rasonable request.

